# Factors Associated with Measles Vaccine Immunogenicity in Children at University Teaching Hospitals, Lusaka, Zambia

**DOI:** 10.1101/2024.10.30.24316424

**Authors:** Priscilla Nkonde Gardner, Cephas Sialubanje, Jimmy Hangoma, Roma Chilengi, Muzala Kapina, Musole Chipoya, Kelvin Mwangilwa, Lillian Lamba, Musaku Mwenechanya, Rodgers Chilyabanyama, Mpanga Kasonde, Soo Young, Davie Simwaba, Isaac Fwemba

**Affiliations:** School of Public Health, University of Zambia^1^, Lusaka, Zambia; Zambia National Public Health Institute, Lusaka, Zambia; Department of Child Health, University Teaching Children’s Hospital, Lusaka, Zambia; School of Public Health, Levy Mwanawasa Medical University, Lusaka, Zambia; Health Programs, Resolve to Save Lives, Lusaka, Zambia; Health Programs, Churches Health Association of Zambia, Lusaka, Zambia; School of Health Sciences, Levy Mwanawasa Medical University, Lusaka, Zambia

**Keywords:** Age, Human Immunodeficiency Virus, Immunogenicity Measles vaccination, Zambia

## Abstract

**Background:** Measles poses a significant global public health challenge, particularly in low-resource settings where vaccination coverage is limited. This study examined factors associated with measles vaccine immunogenicity in children aged 2 to 15 years at the University Teaching Children’s Hospital and the HIV Pediatric Centre of Excellence in Lusaka in Lusaka, Zambia.

**Methods:** This was a comparative analytical cross-sectional study conducted from April to July 2024. Blood samples were collected using a questionnaire that was uploaded in Kobo. Data analysis was conducted in R-studio. Descriptive statistics were conducted to summarize respondent data. Bivariate analysis was computed to test the association between the predictor and outcome variables; multivariate logistic regression analysis was conducted to measure the strength of association using adjusted odds ratios (AORs). The p-value<0.05 was considered significant.

**Results:** Children aged 10 to 15 years were less likely to have retained immunity compared to those aged 2 to 4 years *(AOR = 0*.*270, 95% CI [0*.*114–0*.*618], p = <0*.*002)*. HIV-positive children had lower odds of retaining immunity compared to HIV-Negative children *(AOR = 0*.*290, 95% CI [0*.*137– 0*.*594], p = <0*.*001)*. Children who were breastfed had lower odds of waning immunity *(AOR = 0*.*336; 95% CI [0*.*147–0*.*738], p = 0*.*007)* compared to children who were not breastfed. Residing in Lusaka was associated with lower immunity retention compared to children outside the province; however, this finding requires further investigation with a more representative sample size from areas beyond Lusaka. *(AOR = 0*.*250, 95% CI [0*.*066–0*.*859], p = 0*.*031)*.

**Conclusion:** These findings show that age, breastfeeding and HIV status significantly affect retention of measles immunity in children aged 10-15 years. Younger, HIV-negative and breastfed children show better immunity retention. These results highlight the protective role of breastfeeding and the need for booster doses to enhance immunity retention, especially for older and HIV-infected children.

## Introduction

Measles infections are a significant global public health challenge, particularly in low-resource settings where vaccination coverage and access to healthcare may be limited. (1). Immunization remains the most effective strategy for reducing morbidity and mortality from infectious diseases worldwide. However, the immune response to vaccines can vary considerably due to factors such as age, HIV status, and socio-demographic conditions. (2) (3). Previous research has shown, despite receiving antiretroviral therapy (ART), HIV-infected children often exhibit weaker immune responses to vaccines, including the measles vaccine, compared to their HIV-negative peers (4) . HIV-infected infants who discontinue ART have been shown to be more likely to lose their measles immunity by age 4.5 years compared to those who remained on ART. (5).

Research has demonstrated that the efficacy of the measles vaccine can diminish with age, with younger children generally exhibiting stronger immune responses. (6). For example, a study by Puthanakit in Thailand found that older children had significantly lower rates of measles seropositivity compared to younger children, highlighting the need for age-appropriate vaccination strategies (7). Several factors have been shown to affect the efficacy of measles immunizations such as vaccine storage and cold chain (8). Vaccines are expected to be stored between +2 and +8 °C, but exposure to sub-zero temperatures can damage heat-sensitive vaccines like Measles, Tetanus, DTP, and Hepatitis B(16)(17). Thus, maintaining the cold chain is essential to ensuring vaccine quality and effectiveness. However, vaccine exposure to suboptimal temperatures is a widespread challenge affecting vaccine efficacy in low-resource settings such as Zambia (14) (15).

The co-occurrence of HIV and measles in Zambia raises concerns about the effectiveness of current vaccination programs, especially among HIV-infected children. Current evidence suggests that, despite high vaccination coverage and access to ART, HIV-infected individuals may remain vulnerable (9). This highlights the need for improved and sustained immunization strategies for this population. Supplementary immunization activities (SIAs) have been identified as great opportunities for reducing inequalities in measles vaccine coverage, especially in rural and hard to reach areas. However, studies in Zambia have pointed out challenges in implementing SIAs, especially during the COVID-19 pandemic complicating efforts to achieve optimal measles vaccination coverage in the country (10).

The measles prevention initiative in Zambia seeks to reach 95 percent of children each year through the routine immunization program. The country recommends two doses of a measles-containing vaccine (MCV) to be delivered through the routine program at 9 months and 18 months of age as a minimum optimal number of immunizations (11). However, a study on factors associated with zero-dose vaccination status in Zambia’s Southern Province revealed that gaps in vaccine coverage persisted even after mass vaccination campaigns (12). This gap may present a considerable risk, especially for children living with HIV(13). Disparities in immunity levels between HIV-positive and HIV-negative children and adults in Zambia were noted in a seroprevalence (18), highlighting the importance of tackling systemic issues such as cold chain management and addressing the specific requirements of immunocompromised groups.

Despite these insights, there remains a gap in understanding the effect of age, HIV status, and socio-demographic factors on measles vaccine immunogenicity in sub-Saharan African settings, including Zambia where many children and adolescents are living with HIV. This study examined factors associated with measles vaccine immunogenicity in children between the age of 2 to 15 years at the University Teaching Hospital in Lusaka, Zambia. Insight into these factors is important to inform policy and interventions on measles vaccination programs in Zambia.

## Methods Study design

This comparative analytical cross-sectional study design was conducted from the 2^nd^ of April to the 31^st^ of July 2024.

### Study setting

The study was conducted at the University Teaching Children’s Hospital and the Pediatric Centre of Excellence (PCOE) in Lusaka, Zambia, both of which have a high prevalence of measles and HIV. The hospital recorded over 600 measles cases with about 20 deaths between 2022 and 2023 (19)

### Study population and sampling procedure

The study population consisted of children aged 2 to 15 years who had received at least two doses of the measles vaccine, with participants stratified based on their HIV status. The sample size was calculated using the formula for comparing two proportions, accounting for a 10% non-response rate:

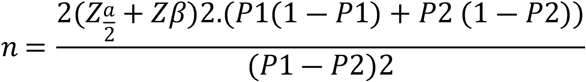

The measles prevalence rate for the infected group (*P*1) was 47.2% (0.472)(20) while the uninfected group (*P*2) had a prevalence rate of 85% (0.85) (21)The significance level was set at 0.05 (95% confidence level), and the power (1−*β*) was 0.80, corresponding to 80% power. The Z-score for *α*/2 (two-sided test) was approximately 1.96, and for *β* (80% power), it was around 0.84. This resulted in a working sample of 98 participants per group, leading to a total of 196 participants, rounded to 200. Data was collected from participants who visited the hospital between April and July using a complete enumeration.

### Inclusion and Exclusion Criteria

To be included in the study, participants needed to be meet the following criteria:

- Age between 2 and 15 years
- Available and complete medical records with immunization history and HIV status,
- Documented measles vaccination status with at least two doses of a measles-containing vaccine
- Confirmed HIV-infection diagnosed according to national guidelines
- HIV-non-infected children without a history of HIV infection or exposure
- Assent from children and consent from their parents or legal guardians

Children below the age of two years and above 15 years were and those with severe immunodeficiency other than HIV, such as primary immunodeficiency disorders were excluded from the study. In addition, children with chronic illnesses or medical conditions that could affect immune response or vaccine efficacy, such as cancer, were also excluded. Further, children who had received blood products or immunoglobulins within the past three months, those currently participating in another clinical trial involving vaccines or immunomodulatory agents, and children with incomplete medical records or missing information about measles vaccination status or HIV diagnosis were excluded. Additionally, children whose parents or guardians declined participation, and those unable to provide assent or cooperate with study procedures due to cognitive or developmental limitations were also excluded.

### Data collection

Kobo a digital data collection tool was used for real-time data entry and storage and enabled the research assistants to gather primary data from both sick and healthy participants who visited the University Teaching Hospital (UTH). Phlebotomists and data collectors made up the field team. These data collectors spoke Nyanja fluently, as did the phlebotomists (blood collectors). They worked under the direction of a team leader and received training in research ethics, biosafety, quality assurance, and the proper use of personal protective equipment. Additionally, the phlebotomists received training on the proper techniques for performing and drawing blood. Supervisors and data collectors received training on how to use the tablet to complete the questionnaire. The field teams also had safety handling procedures and infection prevention and control (IPC) tools. Close supervision, data editing and cleaning, and cross-checking of the surveys’ completeness were further methods used to regulate the quality of the data. Pre-testing the questionnaire in comparable contexts outside the research area allowed for essential revisions to be made to a few of its items. Blood samples of about 2mls were collected from study participants by phlebotomist and health care workers and using the available hospital courier systems, the collected samples were transported to UTH virology laboratory for testing. The testing was conducted in line with the laboratory protocols for measuring measles-specific antibody titres in Zambia. Measles IgG test kits were bought in consultation with the Zambia National Public Health Reference Laboratory. Laboratory quality controls for the test kits were done and were complying. The measles IgG test kits used in this study were OriGene Measles Test Kits accessible at https://cdn.origene.com/datasheet/ea100951.pdf

### Data analysis

Data was cleaned containing Body Mass Index (BMI), HIV status, Place of residence (Lusaka or outside Lusaka), Education status, Age, Sex and immunity status, with a total of 200 participants. Descriptive statistics were performed to observe the characteristics of variables (numbers and percentages were reported as the variables were categorized. Since the condition of the chi-square were met, associations between categorical variables were evaluated using the chi-squared test of independence. Univariable and multivariable logistic regression were employed to identify the variables that predicted waning immunity in the mining community. Using an investigator-led stepwise regression process, multiple logistic regression was utilized to identify the variables impacting waning immunity. To select the variables for the final multiple regression model, all of the predictor variables were first run through the multiple logistic regression command. Then, the predictor variables with the highest p-values were removed from the model one at a time until the predictor variables that best predicted the outcome remained in the model. Ultimately, the best-fit model was selected using the Bayesian information criteria (AIC and BIC) and Akaike’s information criterion for the competing models. The model selected was the one with the lowest AIC and BIC values when compared to the other models. The crude odds ratio (cOR) and adjusted odds ratio (aOR) were displayed together with their respective 95% confidence intervals (CIs). A significance level of 0.05 was assigned to a p-value. Using R statistical analysis software version 4.4.1 data was analyze.

### Ethical consideration

This study was approved by ERES Converge IRB (Ref No. 2024-Feb-025). Permission to conduct the study was granted by the National Health Research Authority (NHRA) (REF: NHREB 008/01/04/2024) and the Ministry of Health. In addition, written consent was obtained from the parents or guardians. The consent forms were written in both English and local languages, including Bemba and Nyanja. The research assistant read and explained the benefits and possible risks of participating in the study to the participants. After gaining full knowledge, participants who agreed to take part were given a written consent form to sign. Respect for persons, beneficence, and justice were upheld throughout the study.

### Participant recruitment algorithm

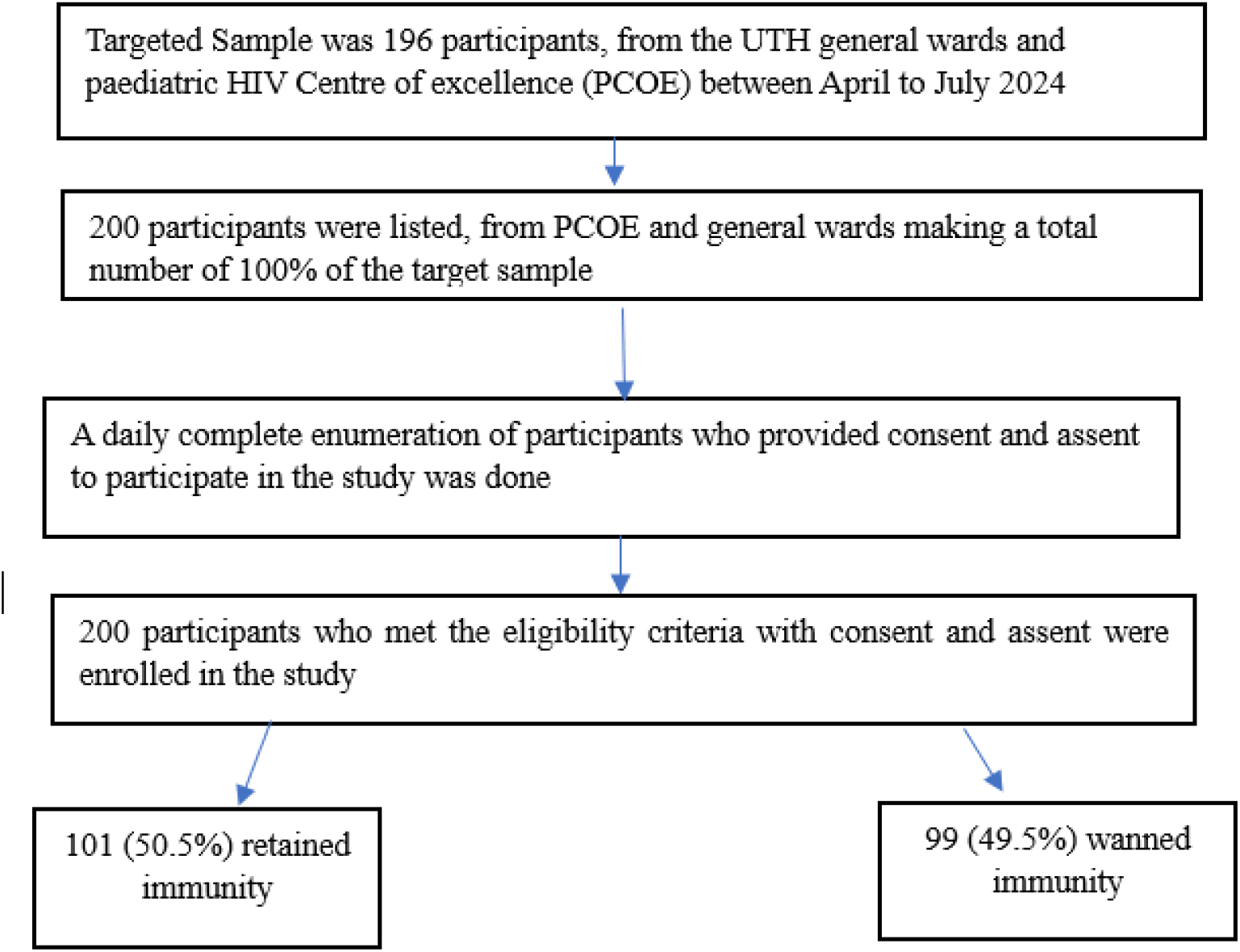

## Results

### Demographic characteristics

Table 1 summarizes the demographic and clinical characteristics of participants, stratified based on HIV status. Almost half of the sample (45%) were aged between 10 and 15 years. And majority (58%) were female and well nourished (75%). Majority (77.5%) were in school and had not received the vaccination on time (51%). Most children (76.0%) were breast fed and resided in Lusaka (93.0%). There was significant difference between the HIV-negative and HIV-positive children about age (p<0.001), BMI (p<0.001), education level (p<0.001), Breastfeeding (p<0.001) and province of residence (p<0.013). No significant difference was found in vaccination timing between the two groups (p = 0.12)

**Table 1:**
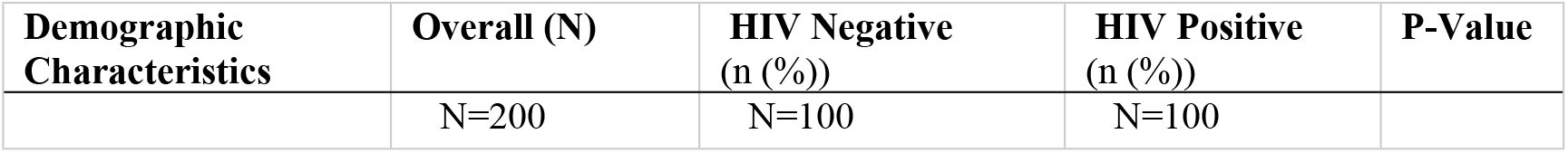
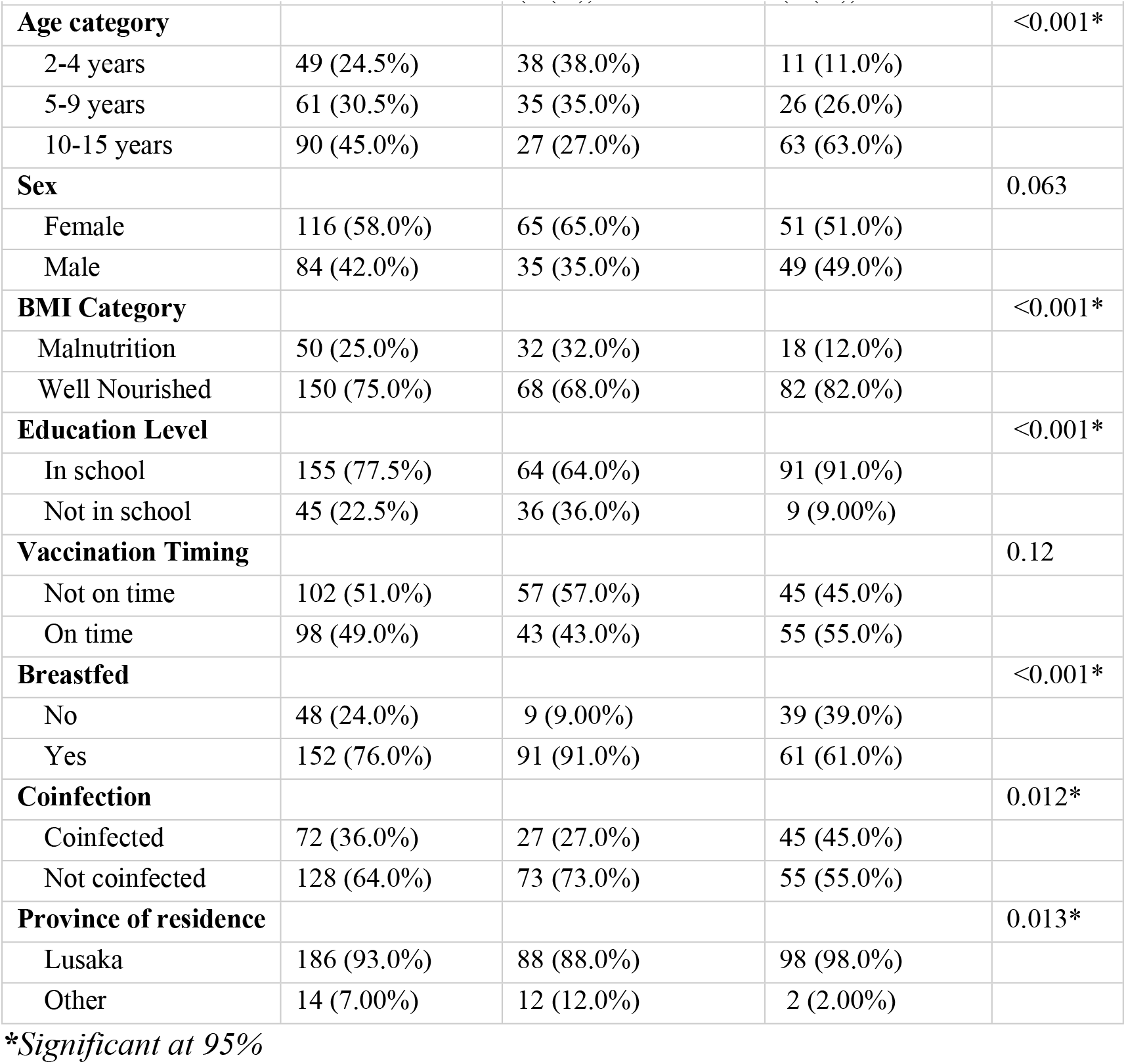
Socio-Demographic Characteristics univariate analysis.

### Unadjusted and Adjusted Multivariate Regression Analysis

Table 2 below shows the results for unadjusted and adjusted multivariate analysis. The adjusted analysis indicates that children aged 10-15 years had lower odds of retaining immunity compared to those aged 2-4 years (AOR= *0*.*228; 95% CI [0*.*076, 0*.*649], p < 0*.*006)*. This means that children in this age group were more likely to lose their immunity compared to the younger age group. For children aged 5-9 years, the AOR was *0*.*854 (95% CI [0*.*300, 2*.*360], p = 0*.*762)*, indicating no significant difference in the odds of waning immunity compared to children aged 2-4 years. Regarding HIV status, children who were HIV-positive had lower odds of retaining their immunity compared to their HIV-negative counterparts (AOR= *0*.*251; (95% CI [0*.*102, 0*.*596], p < 0*.*002)*. This suggests that HIV-positive children were more likely to lose their immunity. Concerning breastfeeding, children who were breastfed had lower odds of waning immunity compared to those who were not breastfed (AOR= *0*.*356; 95% CI [0*.*140, 0*.*873], p = 0*.*0263)*.

**Table 2:** Unadjusted and Adjusted Multivariate Regression Analysis.

| Factors (Reference) | Unadjusted OR | 95% CI | P - value | Adjusted OR | 95%CI | P-Value |
| --- | --- | --- | --- | --- | --- | --- |
| **Age Category** |  |  |  |  |  |  |
| 2-4 years | Ref [1] |  |  | Ref [1] |  |  |
| 5-9 years | 1.105 | 0.502, 2.428 | 0.801 | 0.854 | 0.300, 2.360 | 0.762 |
| 10-15 years | 0.262 | 0.123,0.539 | <0.003 | 0.228 | 0.076, 0.649 | <0.006* |
| **Sex** |  |  |  |  |  |  |
| Females | Ref [1] |  |  | Ref [1] |  |  |
| Male | 1.324 | 0.755, 2.333 | 0.327 | 1.786 | 0.946, 3.830 | 0.075 |
| **BMI Category** |  |  |  |  |  |  |
| Malnourished | Ref [1] |  |  | Ref [1] |  |  |
| Well Nourished | 0.454 | 0.230,0.871 | 0.019* | 0.90 | 0.838,1.080 | 0.474 |
| **Breastfeeding** |  |  |  |  |  |  |
| No | Ref [1] |  |  | Ref [1] |  |  |
| Yes | 0.626 | 0.322,1.201 | 0.250 | 0.356 | 0.140, 0.873 | 0.0263* |
| **Education Status** |  |  |  |  |  |  |
| In school | Ref [1] |  |  | Ref [1] |  |  |
| Not in school | 1.729 | 0.886, 3.439 | 0.112 | 0.714 | 0.245, 2.03 | 0.529 |
| **Vaccination Timing** |  |  |  |  |  |  |
| Not on time | Ref [1] |  |  | Ref [1] |  |  |
| On-time | 0.818 | 0.468,1.424 | 0.478 | 1.050 | 0.440, 2.560 | 0.916 |
| **HIV Status** |  |  |  |  |  |  |
| Negative | Ref [1] |  |  | Ref [1] |  |  |
| Positive | 0.360 | 0.201,0.634 | <0.001 | 0.251 | 0.102, 0.596 | <0.002* |
| **Coinfections** |  |  |  |  |  |  |
| Coinfected | Ref [1] |  |  | Ref [1] |  |  |
| Not coinfectd | 0.808 | 0.497,1.584 | 0.688 | 0.551 | 1.298, 3.127 | 0.111 |
| **Educational Status Caregiver** |  |  |  |  |  |  |
| Never been to school | Ref [1] |  |  | Ref [1] |  |  |
| Primary | 0.421 | 0.153,1.086 | 0.080 | 0.450 | 0.120, 1.570 | 0.220 |
| Secondary | 0.450 | 0.160,1.192 | 0.116 | 0.438 | 0.108, 1.670 | 0.234 |
| Tertiary | 0.480 | 0.167,1.300 | 0.157 | 0.615 | 0.141, 2.570 | 0.509 |
| **Province of Residence** |  |  |  |  |  |  |
| Within Lusaka | Ref [1] |  |  | Ref [1] |  |  |
| Outside Lusaka | 0.543 | 0.161,1.635 | 0.290 | 0.432 | 0.089, 201 | 0.285 |

### Factors Associated with Measles Immunogenicity

Table 3 shows the results of the multivariable analysis for factors associated with measles immunogenicity. After adjusting for confounders, the final model showed that age, HIV status, and history of breastfeeding were significantly associated with measles immunogenicity. Children aged 10-15 years had significantly lower odds of losing retaining immunity compared to those aged 2-4 years (*AOR = 0*.*270; 95% CI [0*.*114, 0*.*618], p = <0*.*002)*. Regarding HIV status, children living with HIV infection had significantly lower odds of retaining immunity compared to those who are HIV-negative (AOR=*0*.*290 (95% CI [0*.*137, 0*.*594], p < 0*.*001)*. Children who were breastfed had lower odds of waning immunity compared to those who were not breastfed, with an (AOR=*0*.*336 (95% CI [0*.*147, 0*.*738], p = 0*.*007)*. Majority of the study participants were Lusaka residents limiting these finding to Lusaka and not other settings.

**Table 3:** Multivariable Analysis.

| Factors (Reference) | OR | 95%CI | P-Value |
| --- | --- | --- | --- |
| **Age Category** |  |  |  |
| 2-4 years | Ref [1] |  |  |
| 5-9 years | 1.15 | 0.493, 2.686 | 0.742 |
| 10-15 years | 0.270 | 0.114, 0.618 | <0.002* |
| **Sex** |  |  |  |
| Females | Ref [1] |  |  |
| Male | 1.798 | 0.944, 3.499 | 0.077 |
| **Breastfeeding** |  |  |  |
| No | Ref [1] |  |  |
| Yes | 0.336 | 0.147, 0.738 | 0.007* |
| **HIV Status** |  |  |  |
| Negative | Ref [1] |  |  |
| Positive | 0.290 | 0.137, 0.594 | <0.001* |
| **Province of Residence** |  |  |  |
| Within Lusaka | Ref [1] |  |  |
| Outside Lusaka | 0.250 | 0.066, 0.859 | 0.031* |

## Discussion

This study examined the factors affecting measles vaccine immunogenicity in children aged 2 to 15 years at the University Teaching Hospital in Lusaka, Zambia. The results showed that age, breastfeeding, HIV status and province of residence were significantly associated with measles immunogenicity. HIV-negative children had higher odds of retaining immunity compared to HIV-positive children, and those who were breast underweight children also revealed higher odds of waning immunity.

Our findings show that age plays a significant role in the immune response to measles vaccination. Older children aged 10–15 years were significantly more likely to lose their immunity compared to younger age groups, particularly those aged 2–9 years. This finding is consistent with various studies which have indicated that younger children, especially those below 5 years of age, show stronger immune responses to measles vaccination due to maternal antibodies by age two, (22)(23)(24). On the other hand, older children (10 years and above) may experience waning measles immunity over time(25)(26). This finding was consistent with our finding in which this study reported several older children with lower odds of retaining measles immunity after vaccination. In addition, literature has also shown that children vaccinated at younger ages may be at risk of waning immunity as they grow older, (27) (Piot et al., 2019).These findings may call for further investigations especially in children who receive booster doses as early as six months.

Another key finding for this study was HIV status and how it affected measles immunity among the HIV infected children. Literature has shown that even with viral suppression, HIV infected children (23)(29) are more vulnerable to severe measles infections (30) (31). Over time, HIV-infected children show diminished vaccine-induced immunity compared to their uninfected counterparts, requiring closer monitoring (32)(33). These findings are consistent with previous studies which reported similar findings (33). The finding highlights the importance of age-appropriate vaccination schedules in maintaining measles immunity over time especially in HIV infected children. Introducing booster doses for HIV infected children at age 10 to 15 would help contribute to strengthened immunity. In Zambia, the routine immunization program aims to reach 95% of children annually with two doses of measles-containing vaccine (MCV) administered at 9 and 18 months.(11). However, for HIV-infected children, additional booster doses or earlier vaccination at 6 months may be necessary to enhance protection and address the potential for waning immunity as they age (32). This is particularly critical in regions with high HIV prevalence, where maternal antibodies may wane earlier, rendering children more susceptible to measles infection (34). Previous studies have shown a strong retention of immunity in older children after receiving booster vaccine as a protective measure (34).

Other factors associated with a retained measles immunity included breast feeding and nutritional status. Children who were breastfed and well-nourished were more likely to retain their immunity compared to those who were not breastfed and malnourished. This finding aligns with the theory that nutritional status can influence immune response and that malnutrition impairs immunity.(35). This finding contrasts previous studies which have reported enhanced immune responses in underweight individuals. These studies suggest that immunity is enhanced due to compensatory mechanisms that upregulate certain immune pathways (36). Further research is needed to explore this complex relationship (37).

Lastly, majority of the study participants were residing within Lusaka province limiting this finding to the urban settings, underscoring the need for similar studies to be conducted in rural areas to provide a more comprehensive understanding of the geographic variability in measles immunity.

Several limitations associated with this study are worth considering. First, the study did not assess the quality of vaccine administration, including adherence to cold chain protocols and proper vaccine handling, which may have affected vaccine efficacy and contributed to waning immunity. The study was conducted in Lusaka district which is predominantly urban, thereby limiting the generalizability of the findings to the wider Zambian population, especially in rural regions where access to healthcare and vaccination practices may vary considerably. Second, the lack of genomic sequencing of measles IgG samples limited the understanding of specific viral strains and mutations that may contribute to waning immunity. Mitigating these limitations in forthcoming research could augment our comprehension of measles immunity and facilitate the formulation of more precise public health interventions.

Despite these limitations, this study has various strengths that substantially enhance the comprehension of measles vaccine immunogenicity in children. First, the broad age range of sampled children facilitated the identification of age-related patterns in diminishing immunity, uncovering essential insights into the variations of immunity throughout different developmental stages. Second, the emphasis on HIV status as a critical factor yielded significant insights into the heightened susceptibility of HIV-positive children, underscoring the necessity for tailored vaccination programs and supplementary booster doses for this population. Third, the study also encompassed factors like breastfeeding and nutritional status, providing a more thorough understanding of broader health determinants and their impact on immune responses. The incorporation of province of residence as a variable underscored potential regional disparity, which is essential for formulating region-specific public health strategies. The alignment of the findings with existing literature supports the credibility of the results, while evidence-based recommendations, including booster vaccinations for HIV-infected children, offer practical guidance for enhancing measles control.

## Conclusion

These findings show that age, breastfeeding and HIV status significantly influence retention of measles immunity in children aged 10-15 years. Living in Lusaka was associated with waning immunity; however, this finding warrants further investigation with a more representative sample size from regions outside Lusaka. Younger children and those without HIV have higher odds of retaining the measles immunity. The findings also demonstrate the potential protective role of breastfeeding and its importance in increasing the likelihood of immunity retention among non-breastfed children. These findings underscore the need for targeted interventions to improve immunity retention, particularly for older children and individuals living with HIV. The findings highlight the importance of introducing booster doses in the immunization schedule for older and HIV-infected children in Zambia to enhance retention of their immunity. Finally, our finding suggests a need for further research with an experimental design, to explore the underlying factors and mechanisms contributing to this measles immunity retention.

## Data Availability

The data supporting the findings of this study are available upon reasonable request. Due to ethical and privacy concerns, the data are not publicly available. However, de-identified data can be provided by contacting the corresponding author with appropriate ethical clearance or institutional approval. The dataset generated and analyzed during the current study is stored at the University Teaching Children’s Hospital, Lusaka, Zambia.

NA

## Acknowledgments

Gratitude is extended to the staff and management of the University Teaching Hospital in Lusaka, the National Virology Laboratory, the ZNPHI Reference Lab, the Pediatric Centre of Excellence, and the University of Zambia for their support and collaboration on this project.

